# CIITA G-286A promoter polymorphism impairs monocytes HLA-DR expression in septic shock and is rescued by interferon-γ

**DOI:** 10.1101/2021.02.19.21252097

**Authors:** Jordi Miatello, Anne-Claire Lukaszewicz, Valérie Faivre, Stéphane Hua, Kim Zita Martinet, Christine Bourgeois, Lluis Quintana-Murci, Didier Payen, Michele Boniotto, Pierre Tissières

**Author notes:** **Corresponding authors:** Pierre Tissières, MD, DSc, Institute of Integrative Biology of the Cell, CNRS, CEA, Paris Saclay University, 1 Avenue de la Terrasse, 91190 Gif-sur-Yvette, France. Both authors contributed equally (Michele Boniotto, Pierre Tissières). **Statement of equal author contributions:** MB and PT contributed equally. **Statement of article series:** none. **Statement of prior presentation:** presented partially in abstract form at the Cell symposia: Human immunity, Lisbon, Portugal, August 2012.

## Abstract

Monocyte HLA-DR is an increasingly recognized markers of sepsis-induced immunodepression, but its regulatory mechanisms remain poorly understood in sepsis. Several evidence for positive selection on the 5’ promoter region of HLA class II transactivator (*CIITA*) gene, the master regulator of MHC class II, have been gathered in the European population, and its role in sepsis has never been demonstrated, whilst suggested in autoimmune disease. We aim to describe the effect of rs3087456 polymorphism, localized on *CIITA* promoter III (pIII), on mortality of patients with septic shock, and investigate the mechanisms regulating HLA-DR expression. Genotyping of 203 patients with septic shock showed that, in A dominant model, GG genotype was associated with 28-day mortality (OR 2.29; 95%CI: 1.01 to 5.22; *P* = 0.043). Monocyte HLA-DR remained low in patients with GG genotype whereas it increases as early as at the end of the first week in intensive care in patients with AA or AG genotype. Using site-directed mutagenesis, *in vitro* reporter gene promoter activity of the pIII was decreased in GG genotype in monocyte cell line. Interferon-γ (IFN-γ) restored pIII activity in GG genotype as well as restore, in *ex vivo* experiment in healthy volunteers, *CIITA* pIII expression of GG genotype. Hereby, we demonstrated that rs3087456, a positively selected polymorphism of *CIITA* proximal promoter, significantly impact monocyte HLA-DR expression in patients with septic shock through CIITA promoter activity, that can be rescued using IFN-γ, offering a new perspective in genetic susceptibility to sepsis and targeted immunomodulatory therapy.

**Keypoints:**

- CIITA G-286A polymorphism reduces promotor activity and significantly impact monocyte HLA-DR expression and mortality in septic shock
- Downregulatory effects of CIITA G-286A polymorphism on monocyte HLA-DR expression can be reverse by IFN-γ in patients with septic shock

## INTRODUCTION

Sepsis is a life-threatening condition with more than 19 million cases and 5 million deaths per year worldwide and is recognized as a global priority by the World Health Organization ^1,2^. Incidence of sepsis has increased during last decades because of advancing age, immunodepression and multidrug-resistant infections ^3^. Nevertheless, mortality remains stable pending the prompt administration of antibiotics, the use of fluid resuscitation and vasopressors to reverse hypotension and maintain tissue perfusion^4^. Sepsis is now widely recognized as an inappropriate host response to infection with transient or persistent state of immunodepression ^5,6^. Diagnosis and specific therapies targeting host response, such as anti-endotoxin serum or purified anti-endotoxin ^7^, high-dose steroids or inhibitors of the pro-inflammatory cytokines IL-1, IL-1 receptor, and TNF-α ^8–10^, anticoagulants ^11,12^ (activated protein C) were investigated. Beside strong physiopathologic rationale, most of these studies failed to be beneficial. However, when patients are stratified according to specific immunological phenotypes, these therapies were shown to improve survival, whilst worsening the conditions of others ^13^.

Sepsis-induced immunodepression is now recognized as being associated with both mortality and emergence of secondary infections ^14,15^. To assess the patients’ immunodepression status, use of HLA-DR expression in circulating monocytes (mHLA-DR) is widely recognized ^16^. A persistent low level of mHLA-DR measured by flow cytometry is associated with the development of secondary infections and impaired clinical outcome ^14,15^. Similarly, decreased mRNA levels of several genes controlling HLA-DR protein structure, transport and peptide loading have been associated to poor sepsis outcome ^17–19^. Among these, expression of the master regulator of HLA class II, class II transactivator (*CIITA*), was shown to be reduced in deceased septic patients ^17^. MHC class II expression is controlled at the level of transcription by a highly conserved regulatory module that is found in the promoters of the gene encoding the α-chain and β-chain of all MHC class II molecules (HLA-DP, -DQ, -DR, HLA-DM and -DO). *CIITA* gene expression is regulated mainly at the level of transcription that is driven by a large regulatory region containing four distinct promoters. Among these four promoters regions, three main *CIITA* isoforms are produced but their functions are not clear yet. Cell specificity is determined by transcriptional control of the *CIITA* gene through the differential activity of three of the four promoter regions. Promoter I (pI) is used mainly in dendritic cells and is - induced IFN-γ in macrophages, pIII is activated in B cells and activated T cells, and pIV is induced by IFN-γ in cells that do not express usually the MHC class II ^20^. Mutations of *CIITA* gene are causing severe haplotype insufficiency, such in bare lymphocytes syndrome, that impairs expression of MHC class II genes ^21,22^. Among the various single nucleotide polymorphism (SNP) described, rs3087456 in the *CIITA* pIII is associated to a variety of immune-inflammatory disease and cancer ^23,24^. Swanberg et al ^23^ showed that rs3087456 influence CIITA and HLA-DR transcripts in human peripheral blood mononuclear cells (PBMCs) stimulated by IFN-γ and demonstrate its association with rheumatoid arthritis, multiple sclerosis, and myocardial infarction in Swedish population. Other association of this SNP with autoimmune diseases such as systemic lupus erythematosus ^25^, myasthenia gravis ^26^, celiac disease ^27^ and primary adrenal insufficiency ^28^ were published. However, this association is debated suggesting that gene-gene, gene-environment may interfere. As an example, a first meta-analysis found no significant association between rs3087456 and rheumatoid arthritis, whilst a significant association, further increased in Scandinavian cohorts, was identified in other study suggesting that population genetics may play an important role in rs3087456 associated clinical phenotypes ^29,30^. Vasseur et al ^31^ provided some evidence of a positive selection of the 5’ genomic region of *CIITA*. The positive selection, detected by LD-based tests and Fst tests, identified as highly differentiated in Europe (27 % have a G variant, 73 % an A). In addition, SNP rs12596540 is shown to be in high linkage disequilibrium (r^2^ >0.6) with SNP rs3087456 ^31^. Considering the role of CIITA in the regulation of HLA-DR, and its association with immune-inflammatory diseases, we hypothesize that population-selected rs3087456 or its linked SNP rs12596540, may impact monocyte HLA-DR expression and sepsis outcome in critically septic patient, and evaluate the role of IFN-γ in reversing the phenotype.

## MATERIALS AND METHODS

### Study Population and Human Samples

Two hundred and twenty-one DNA samples were obtained from septic shock patients admitted in the Critical Care Unit of the department of Anesthesiology, Lariboisière University Hospital, Paris, France, between February 2004 and November 2005. The study was approved by the Cochin Hospital Ethics Committee (# CCPPRB 2061), Assistance Publique Hôpitaux de Paris. Of the 221 samples, 17 had insufficient amount of DNA for SNP genotyping and one patient had no data for the primary outcome. All patients fulfilled criteria of the American College of Chest Physicians/Society of Critical Care Medicine for septic shock ^32^. Patients were followed up throughout their ICU stay. Only subjects with clinical and biological data available for more than 7 days were analyzed. Clinical and biological characteristics were also collected. These included demographic characteristics (e.g. age, gender), biological data such as measurement of mHLA-DR on a twice weekly base, severity scores (e.g. SAPS II: Simplified Acute Physiology Score II and SOFA: Sequential Organ Failure Assessment), infection foci and complete blood cell count. For *ex-vivo* and *in-vitro* experiments, peripheral blood from healthy blood donors were collected at the French national blood collection center (*Etablissement Francais du Sang, EFS*). According to EFS standardized procedures for blood donation, written informed consent was obtained from healthy volunteers.

### SNP Genotyping

Genomic DNA (gDNA) was extracted from 200 µL of peripheral whole blood using the NucleoSpin Blood kit (Macherey-Nagel, Düren, Deutschland) following manufacturer’s instruction. Genomic DNA concentration was measured (NanoDrop 2000 spectrophotometer, ThermoFisher, Waltham, MA) and quality assessed (260/280 ratio between 1.7-1.9). Presence of two single nucleotide polymorphisms −286G/A (rs3087456) and −712G/A (rs12596540) located on the *CIITA* promoter III were analyzed. Genotyping was performed blinded to the clinical data. SNP genotype was performed using commercially available TaqMan assays (Life Technologies, La Jolla, CA). Probe and primer combinations were designed to discriminate the two SNPs (Supplemental material, Table 1). Real time allelic discrimination assays were performed using SsoFast Probes Supermix (Bio-Rad, Hercules, CA) and a CFX-96 real time PCR system (Bio-Rad, Hercules, CA).

**Table 1.**
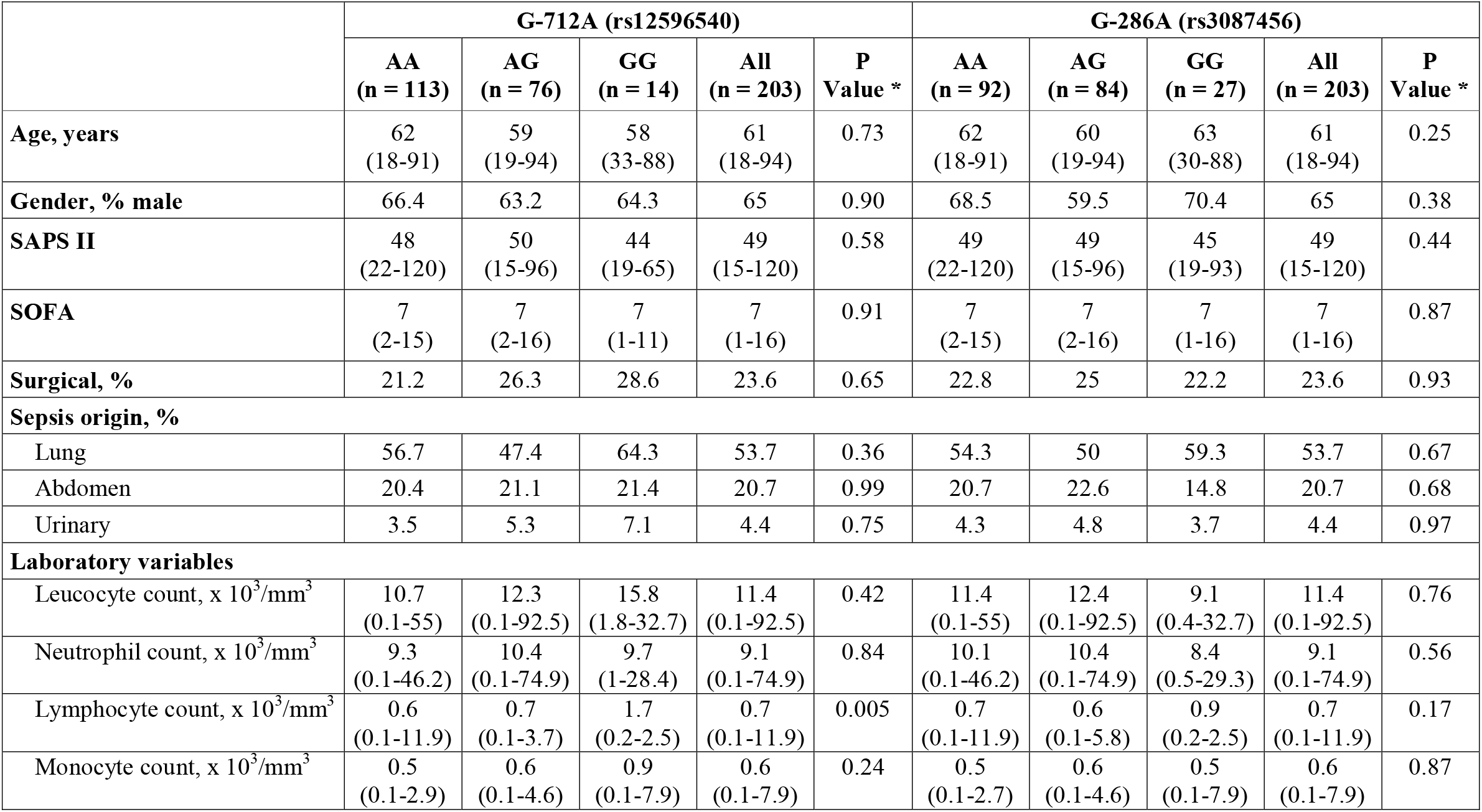
Baseline characteristics of patients in septic shock. SAPS II: Simplified Acute Physiology Score II; SOFA = Sequential Organ Failure Assessment. Data are median (interquartile range) for continuous variable.*P values were calculated with the use of chi-square test and Kruskal-Wallis test.

|  | **G-712A (rs12596540)** |  |  |  |  | **G-286A (rs3087456)** |  |  |  |  |
| --- | --- | --- | --- | --- | --- | --- | --- | --- | --- | --- |
|  | **AA**<br>**(n = 113)** | **AG**<br>**(n = 76)** | **GG**<br>**(n = 14)** | **All**<br>**(n = 203)** | **P**<br>**Value \*** | **AA**<br>**(n = 92)** | **AG**<br>**(n = 84)** | **GG**<br>**(n = 27)** | **All**<br>**(n = 203)** | **P**<br>**Value \*** |
| **Age, years** | 62<br>(18-91) | 59<br>(19-94) | 58<br>(33-88) | 61<br>(18-94) | 0.73 | 62<br>(18-91) | 60<br>(19-94) | 63<br>(30-88) | 61<br>(18-94) | 0.25 |
| **Gender, % male** | 66.4 | 63.2 | 64.3 | 65 | 0.90 | 68.5 | 59.5 | 70.4 | 65 | 0.38 |
| **SAPS II** | 48<br>(22-120) | 50<br>(15-96) | 44<br>(19-65) | 49<br>(15-120) | 0.58 | 49<br>(22-120) | 49<br>(15-96) | 45<br>(19-93) | 49<br>(15-120) | 0.44 |
| **SOFA** | 7<br>(2-15) | 7<br>(2-16) | 7<br>(1-11) | 7<br>(1-16) | 0.91 | 7<br>(2-15) | 7<br>(2-16) | 7<br>(1-16) | 7<br>(1-16) | 0.87 |
| **Surgical, %** | 21.2 | 26.3 | 28.6 | 23.6 | 0.65 | 22.8 | 25 | 22.2 | 23.6 | 0.93 |
| **Sepsis origin, %** |  |  |  |  |  |  |  |  |  |  |
| Lung | 56.7 | 47.4 | 64.3 | 53.7 | 0.36 | 54.3 | 50 | 59.3 | 53.7 | 0.67 |
| Abdomen | 20.4 | 21.1 | 21.4 | 20.7 | 0.99 | 20.7 | 22.6 | 14.8 | 20.7 | 0.68 |
| Urinary | 3.5 | 5.3 | 7.1 | 4.4 | 0.75 | 4.3 | 4.8 | 3.7 | 4.4 | 0.97 |
| **Laboratory variables** |  |  |  |  |  |  |  |  |  |  |
| Leucocyte count, x 10<sup>3</sup>/mm<sup>3</sup> | 10.7<br>(0.1-55) | 12.3<br>(0.1-92.5) | 15.8<br>(1.8-32.7) | 11.4<br>(0.1-92.5) | 0.42 | 11.4<br>(0.1-55) | 12.4<br>(0.1-92.5) | 9.1<br>(0.4-32.7) | 11.4<br>(0.1-92.5) | 0.76 |
| Neutrophil count, x 10<sup>3</sup>/mm<sup>3</sup> | 9.3<br>(0.1-46.2) | 10.4<br>(0.1-74.9) | 9.7<br>(1-28.4) | 9.1<br>(0.1-74.9) | 0.84 | 10.1<br>(0.1-46.2) | 10.4<br>(0.1-74.9) | 8.4<br>(0.5-29.3) | 9.1<br>(0.1-74.9) | 0.56 |
| Lymphocyte count, x 10<sup>3</sup>/mm<sup>3</sup> | 0.6<br>(0.1-11.9) | 0.7<br>(0.1-3.7) | 1.7<br>(0.2-2.5) | 0.7<br>(0.1-11.9) | 0.005 | 0.7<br>(0.1-11.9) | 0.6<br>(0.1-5.8) | 0.9<br>(0.2-2.5) | 0.7<br>(0.1-11.9) | 0.17 |
| Monocyte count, x 10<sup>3</sup>/mm<sup>3</sup> | 0.5<br>(0.1-2.9) | 0.6<br>(0.1-4.6) | 0.9<br>(0.1-7.9) | 0.6<br>(0.1-7.9) | 0.24 | 0.5<br>(0.1-2.7) | 0.6<br>(0.1-4.6) | 0.5<br>(0.1-7.9) | 0.6<br>(0.1-7.9) | 0.87 |

### Cell isolation and culture

Peripheral blood mononuclear cells from healthy volunteers were isolated by Ficoll density-gradient centrifugation. B cells from blood donors were isolated using the MS columns and the CD19 Microbeads kit (Miltenyi Biotec, Bergisch Gladbach, Germany) following manufacturer’s protocols. Monocytes were isolated by positive selection using MS columns and the CD14 Microbeads kit (Miltenyi Biotec, Bergisch Gladbach, Germany). To assess the cell purity by flow cytometry analysis, B cells were stained with fluorescein isothiocyanate (FITC)-conjugated anti-CD20 (Miltenyi Biotec, Bergisch Gladbach, Germany), and monocytes were labeled with phycoerythrin (PE)-conjugated anti-CD14 (BD Biosciences, Le Pont de Claix, France). Isolated cells had an average purity of 90% (± 8%) as determined by flow cytometry analysis.

PBMCs were cultured in 24-well plates at 2 × 10^6^ cells/ml in RPMI with 10% FBS at 37°C, 5% CO_2_. Isolated CD14^+^ cells were cultured at 2.5 × 10^5^ in 24-well plates in DMEM with 5% FBS at 37°C, 5% CO_2_. Isolated CD19^+^ cells were cultured at 10^5^ cells in 96-well plates in RPMI 10% FBS at 37°C, 5% CO_2_. Cultured cells were incubated in the presence or absence of 50 UI/ml of human recombinant IFN-γ (ImmunoTools, Friesoythe, Germany). After 6 hours, CD14^+^ and CD19^+^ cells were harvested for RNA extraction, whilst after 24 hours, PBMCs were collected for FACS analysis.

### Total RNA isolation and quantitative PCR analysis

Total RNA was isolated from monocytes and B cells using the NucleoSpin® RNA XS (Macherey-Nagel, Düren, Deutschland), following manufacturer’s instruction. cDNA was generated from 200 ng of total RNA using iScript™ cDNA Synthesis Kit (Bio-Rad, Hercules, CA). Primers for each CIITA isoform and for the large subunit of ribosomal protein (RPLPO), were designed using the software Beacon Designer 7.0 (PREMIER Biosoft, Palo Alto, CA) and are reported in together with working concentrations and amplification efficiencies (Supplemental material, Table 1). RPLPO was used as a reference gene and the relative quantity of the transcript was calculated by the 2^-ΔΔCt^ method ^33^. PCR reactions were performed in a CFX96 real time PCR system (Bio-Rad Hercules, CA) using the SsoFast EvaGreen Supermix (Bio-Rad, Hercules, CA).

### Flow cytometry analysis

For septic patients, mHLA-DR analysis was described elsewhere ^34^. Shortly, peripheral blood was sampled in EDTA anticoagulants tubes at different time points (day 1, 2, 7, 14, 21 and 28) following admission. Quantification of mHLA-DR on circulating monocytes was performed using a standardized flow cytometry assay as previously described ^34,35^. To quantify, the median fluorescence intensity of the entire monocyte population was then transformed to number of antibodies per cell (AB/C) using calibrated PE beads (BD QuantiBRITE™ PE BEADS, Becton-Dickinson San Jose, CA).

For analysis of healthy donors’ blood, isolated peripheral blood mononuclear cells (PBMCs), monocytes and B cells were washed once in PBS. After being washed once in cold PBS, cells were incubated at 4°C for 15’ with the Human Fc Receptor Binding Inhibitor (eBioscience, San Diego, CA) to avoid unspecific binding of labeled antibodies. Cells were then incubated with fluorescently labeled antibodies were added for 15 min at 4°C. The antibodies mix contained: Fluorescein isothiocyanate (FITC)-conjugated anti-CD20 (mouse monoclonal IgG1κ, clone LT20, Miltenyi Biotec, Bergisch Gladbach, Germany), phycoerythrin (PE)-conjugated anti-CD14 (mouse monoclonal IgG2α,κ, clone M5E2, BD Biosciences, Franklin Lakes NJ) and PE-cyanine 7 (PE-Cy7)-conjugated anti-HLADR (mouse monoclonal IgG2α,κ, clone G46-6, BD Biosciences, Franklin Lakes, NJ). After two PBS washes, cells were suspended in 1% paraformaldehyde. Flow cytometry was performed on a BD FACSCanto (BD Biosciences, Franklin Lakes, NJ) and results analyzed by the software FlowJo (TreeStar Inc, Ashland, OR).

### Cell culture and Transfection

Human leukemic monocyte lymphoma cell line U937 and human Burkitt’s lymphoma cells (Raji) were cultured in RPMI 1640 containing 10% heat-inactivated foetal bovine serum (FBS), 1.0 mM sodium pyruvate, amino acids, 20 mM HEPES, 100 U/ml penicillin, 100 μg/ml streptomycin at 37°C in 5% CO_2_. 4 × 10^5^ cells were transfected with 1 µg of reporter plasmid and 10 ηg of pRL Renilla luciferase vector (Promega, Madison, WI) using Turbofect™ Transfection Reagent (Thermo Fisher Scientific, Fermentas, Waltham, MA). After transfection, the cells were incubated for 30 hours and stimulated by 50 UI/ml rIFN-γ (ImmunoTools, Friesoythe, Germany) for 6h or left untreated.

### Plasmid and Dual-Luciferase Reporter assays

The 1200 bp of the type III promoter of MHCIITA was cloned in pGL-3 firefly luciferase reporter vector (Promega, Madison, WI). Single-nucleotide polymorphisms G-286A (rs3087456) and G-712A (rs12596540) were inserted by using site-specific mutagenesis following published protocols. All the inserts were verified by Sanger sequencing. Dual-Luciferase assay was performed with the Dual-Glo luciferase Assay (Promega, Madison, WI), following manufacturer’s instructions. Firefly and Renilla luciferase substrates fluorescence was recorded by a TriStar LB 941 (Berthold Technologies, Wilbad, Germany). As an internal control, pRL-TK, expressing the Renilla luciferase under the control of the constitutive TK promoter was co-transfected (Promega, Madison, WI). Results are represented by the mean value and the standard deviation and are expressed as relative luciferase activity (ratio of firefly luciferase activity to renilla luciferase activity). To confirm the results, experiments were performed in triplicates and each one was carried out three times.

### Statistical analysis

Allele and genotype frequencies were calculated by direct counting. Hardy-Weinberg equilibrium for the two SNPs was assessed by the software Arlequin v3.11. Analysis of association between genotype and binomial clinical outcomes using different genetic models was performed by using the software plink (http://pngu.mgh.harvard.edu/~purcell/plink/). Results are presented as range and median for continuous variables and as proportion of total for categorical variables. Differences in HLA-DR, SOFA and SAPS II scores for the different genotypes were analyzed by using the Kruskal-Wallis test run in STATA software. The Wilcoxon rank sum test was applied to analyze continuous variables between different genotypes. Mann-Whitney Test was applied for analysis of luciferase experiments by using the software GraphPad Prism 8.0. *P* values < 0.05 were considered significant.

## RESULTS

### Severity and mortality in septic shock in association to *CIITA* rs12596540 and rs3087456 genotypes

Two hundred and three patients with septic shock were included in the analysis. The main patient characteristics are shown in Table 1. Allele frequency and genotype distribution were similar to that observed by Vasseur et al ^31^: rs3087456: AA 45%, AG 41%, GG 13%; rs12596540: AA 56%, AG 37%, GG 7%. Most cell counts were not significantly different between genotypes, but patients who had the *CIITA* rs12596540 GG genotype had a higher lymphocyte count. Both polymorphisms met the Hardy Weinberg law in our cohort. To determine whether *CIITA* rs12596540 and rs3087456 SNPs were associated with severity and mortality in septic shock, genotype frequencies were determined. As reported in Table 1, no significant association was found for both SNPs with clinical severity scores (SAPS II and SOFA scores at day 1). As given in Table 2, we observed a trend of increasing mortality in patients with GG genotype for both SNPs. In an A-dominant model, patients homozygous for allele *CIITA* rs3087456 GG genotype had a significant increased risk of 28-day mortality (55.6% for GG genotype vs. 35.2% for AA and AG genotypes, OR: 2.298, CI 95% [1.013-5.217], *P = 0*.*043*) (Table 2). The presence of rs3087456*A allele was associated with a decreased risk of death from septic shock. We noted that there was a non-significant trend towards an increase in 7-day mortality for GG genotype for rs12596540 (AA or AG: 25.9% vs. GG: 50%, OR: 2.857, CI 95% [0.954-8,558], *P = 0*.*05*). No association was found for secondary infections between the different genotypes of CIITA promoter III gene in both SNPs (Table 2**Error! Reference source not found**.). We further evaluate the impact of both SNPs on mHLA-DR expression in septic shock patients.

**Table 2.** Genotyped related mortality. *P values were calculated with the use of chi-square test and Kruskal-Wallis test.

|  | **G-712A (rs12596540)** |  |  |  |  | **G-286A (rs3087456)** |  |  |  |  |
| --- | --- | --- | --- | --- | --- | --- | --- | --- | --- | --- |
|  | **AA**<br>**(n = 113)** | **AG**<br>**(n = 76)** | **GG**<br>**(n = 14)** | **All**<br>**(n = 203)** | **P**<br>**Value \*** | **AA**<br>**(n = 92)** | **AG**<br>**(n = 84)** | **GG**<br>**(n = 27)** | **All**<br>**(n = 203)** | **P**<br>**Value \*** |
| **28-d mortality, n (%)** | 45 (39.8) | 24 (31.6) | 8 (57.1) | 77 (37.9) | 0.16 | 35 (38) | 27 (32.1) | 15 (55.6) | 77 (37.9) | 0.10 |
| **7-d mortality, n (%)** | 31 (27.2) | 18 (23.7) | 7 (50) | 56 (27.6) | 0.13 | 25 (27.2) | 21 (25) | 10 (37) | 56 (27.6) | 0.47 |
| **Secondary infections, n (%)** | 23 (20.3) | 18 (23.7) | 2 (14.3) | 43 (21.2) | 0.70 | 19 (20.7) | 20 (23.8) | 4 (14.8) | 43 (21.2) | 0.61 |

**A-dominant model**
|  | **G-712A (rs12596540)** |  |  |  |  | **G-286A (rs3087456)** |  |  |  |  |
| --- | --- | --- | --- | --- | --- | --- | --- | --- | --- | --- |
|  | **AA or AG**<br>**(n = 189)** | **GG**<br>**(n = 14)** | **OR** | **95% CI** | **P**<br>**Value \*** | **AA or AG**<br>**(n = 189)** | **GG**<br>**(n = 27)** | **OR** | **95% CI** | **P**<br>**Value \*** |
| **28-d mortality, n (%)** | 69 (36.5) | 8 (57.1) | 2.319 | 0.773-6.960 | 0.13 | 62 (35.2) | 15 (55.6) | 2.298 | 1.013-5.217 | 0.043 |
| **7-d mortality, n (%)** | 49 (25.9) | 7 (50) | 2.857 | 0.954-8.558 | 0.05 | 46 (50) | 10 (37) | 1.662 | 0.710-3.891 | 0.24 |
| **Secondary infections, n (%)** | 41 (21.7) | 2 (14.3) | 0.602 | 0.129-2.796 | 0.49 | 39 (20.7) | 4 (14.8) | 0.611 | 0.199-1.872 | 0.38 |

### *CIITA* rs12596540 and rs3087456 genotypes and mHLA-DR surface expression

In septic shock patients, circulating mHLA-DR expression was measured at different time point during the ICU stay. For the rest of the analysis, due to the mortality data suggesting an effect only in an A-dominant model, we considered only this later model. For rs12596540 and rs3087456, septic patients with AA or AG genotype showed a significant early increase in mHLA-DR level, whereas patients with GG genotype displayed a persistent low mHLA-DR values (Wilcoxon signed rank test, *P < 0*.*03* vs. basal) (Figure 1**Error! Reference source not found**.).

**Figure 1.**
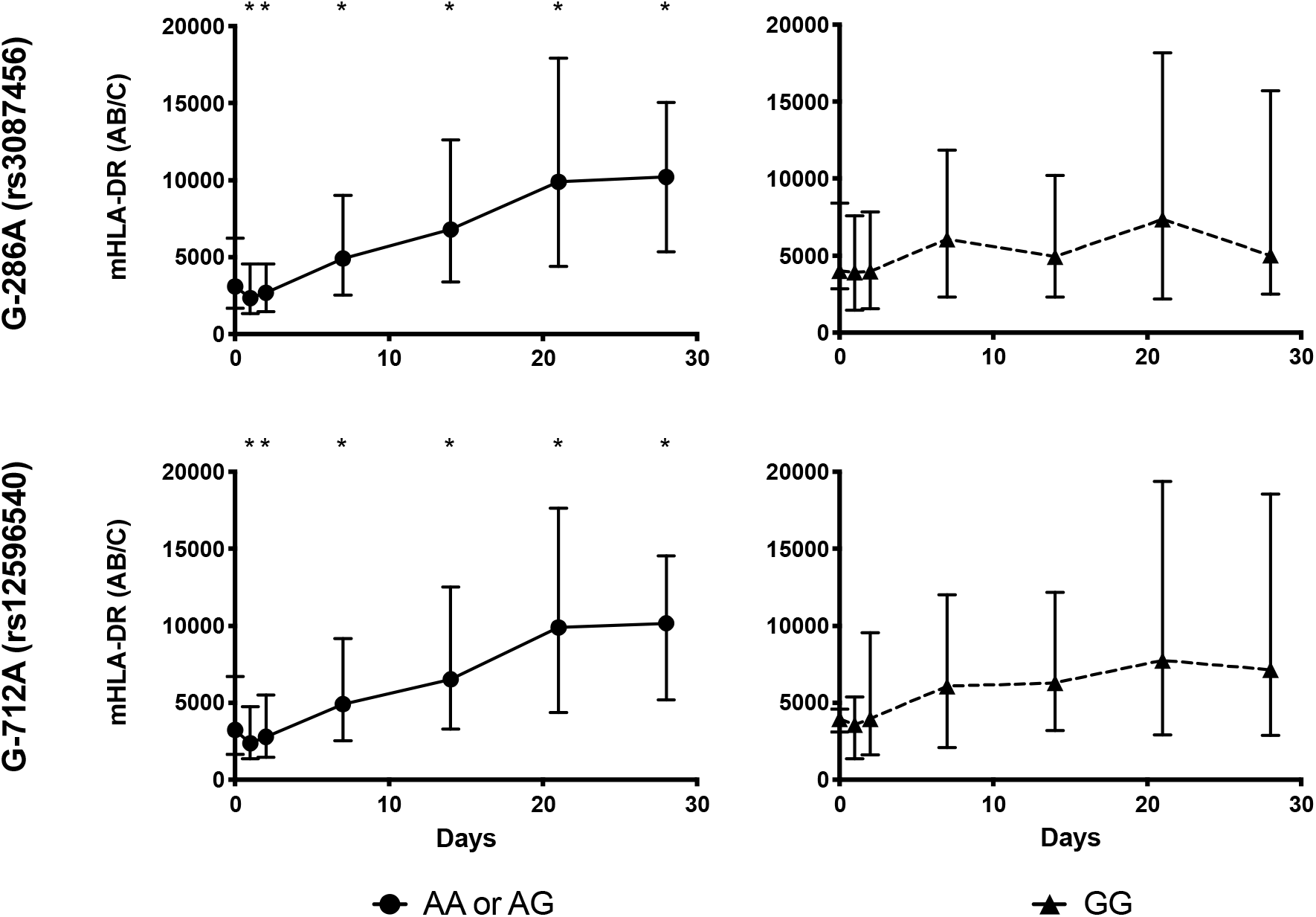
Monocyte HLA-DR expression in 203 patients with septic shock. Circulating monocyte HLA-DR expression is expressed as number of antibodies bound per cell (AB/C) as defined using calibrated PE beads (BD QuantiBRITE™ PE BEADS, Becton-Dickinson San Jose, CA). Results are presented as median with 95% CI and by genotype of CIITA rs12596540 and rs3087456 in a dominant model.*P < 0.03 vs. basal (Wilcoxon signed rank test).

To investigate the role of *CIITA* polymorphisms in healthy individuals, we measured basal mHLA-DR expression in 50 controls and evaluate its response to IFN-C *ex-vivo* stimulation. There was no difference between different genotypes in both polymorphisms (rs3087456: AA or AG = 1371 [95%CI, 1154 to 1625] MFI vs. GG = 1192 [95% CI, 864 to 2207] MFI, *P* = 0.87; rs12596540: AA or AG = 1374 [95%CI, 1154 to 1642] MFI vs. GG = 1137 [95%CI, 864 to 1889] MFI, *P =* 0.45; Figure 2). Following IFN-γ stimulation, mHLA-DR significantly increased in monocytes of healthy volunteers with AA or AG genotype (rs3087456: 1455 vs. 3569 MFI [CI 95%, 1572 to 2656], *P* < 0.0001; rs12596540: 1471 vs. 3518 MFI [CI 95%, 1506 to 2587], *P* < 0.0001; Figure 2). For rs3087456, mHLA-DR was significantly lower for GG genotype after stimulation with IFN-γ (AA or AG = 3919 [95%CI, 2912 to 4434] MFI vs. GG = 1898 [95% CI, 1284 to 2835] MFI, *P* < 0.05, Figure 2). In healthy volunteers, a homozygous GG status did not allow such a high response to IFN-γ. As with mortality data, we did not observe significant difference in mHLA-DR expression with rs12596540 genotypes and focused the rest of the analysis on rs3087456 genotype. To evaluate the effect of rs3087456 on *CIITA* expression regulation, we tested its specific activity on pIII on different leukemoid cell lines.

**Figure 2.**
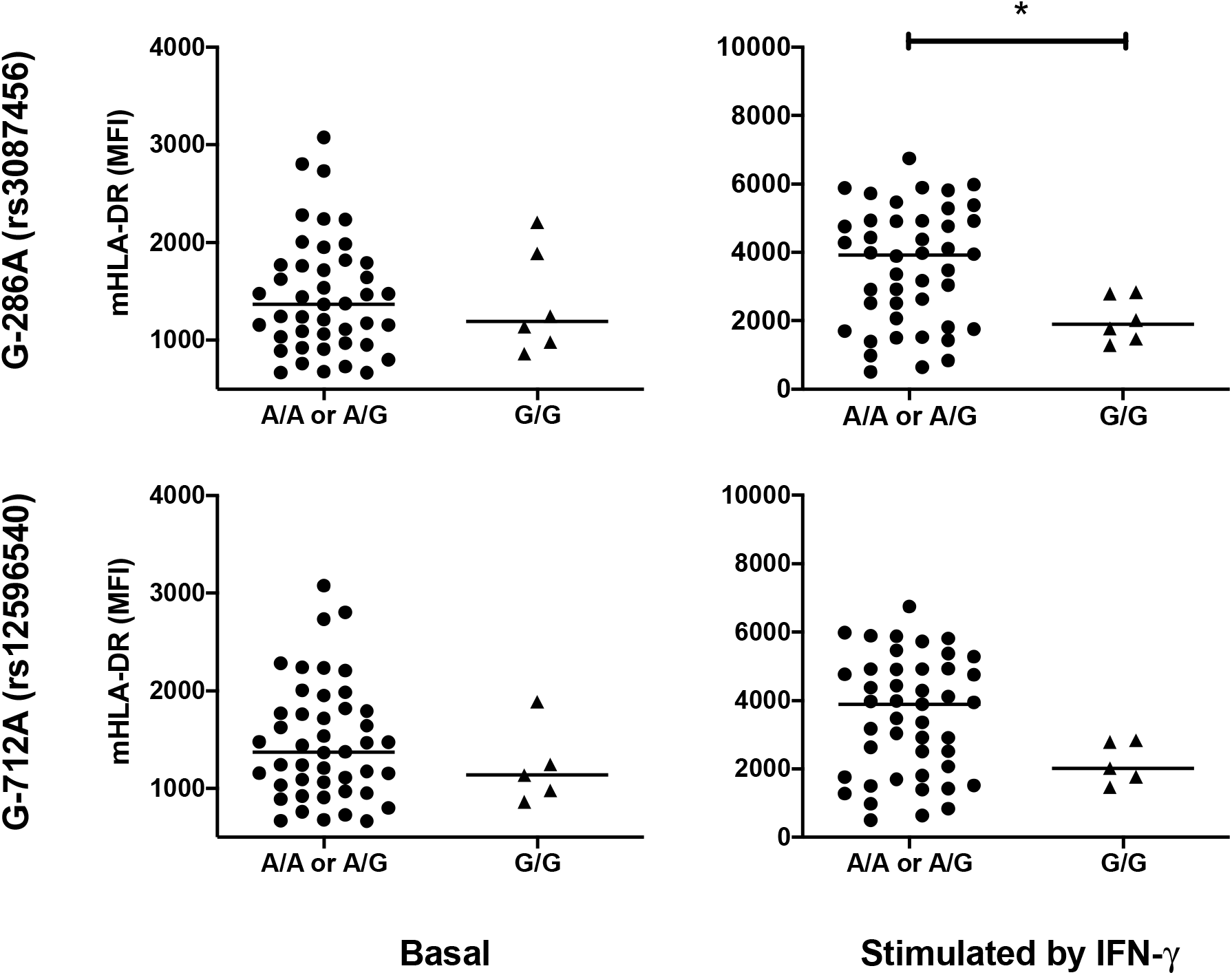
HLA-DR expression in PBMCs of healthy volunteers. Mean fluorescence intensity (MFI) are presented as median with 95% CI and by genotype of CIITA rs12596540 and rs3087456 in a dominant model. P values were calculated using Mann-Whitney Test.*P < 0.05.

### Promoter activity of A/G at position −286 (rs3087456) in the type III promoter of *CIITA*

Functional consequence of this polymorphism on pIII activity was evaluated using reporter gene assays. To understand the mechanism by which *CIITA* gene transcripts are expressed, dual luciferase reporter assays were used to detect specific *CIITA* promoter activity. After transfection into a monocyte-like cell line (U937), G-286A*rs3087456 pIII activity significantly decreased compared with the activity of wild-type (WT) construct (*P* < 0.001) (Figure 3). After stimulation by IFN-γ, activity of G-286A*rs3087456 pIII recovered a similar activity than WT promoter (*P* < 0.001) (Figure 3). As isoforms of pIII are specifics of B and activated-T cells, we transfected B lymphocyte-like cell line (Raji) and T lymphocyte-like cell line (Jurkat T). In B cells, luciferase activity of G-286A*rs3087456 pIII significantly decreased compared with WT (*P* < 0.05), and IFN-γ did not restore activity of mutated pIII (*P*= 0.23) (Figure 3). For T cells, activity of G-286A*rs3087456 pIII failed to show any difference compared to WT promoter both on basal and IFN-γ stimulated condition (Figure 3). These results confirmed that *CIITA* pIII activity is involved in the decrease of mHLA-DR expression in homozygote GG healthy volunteers. To prove the transcriptional effect of G-286A*rs3087456 pIII on *CIITA* expression, we quantified pIII related transcripts.

**Figure 3.**
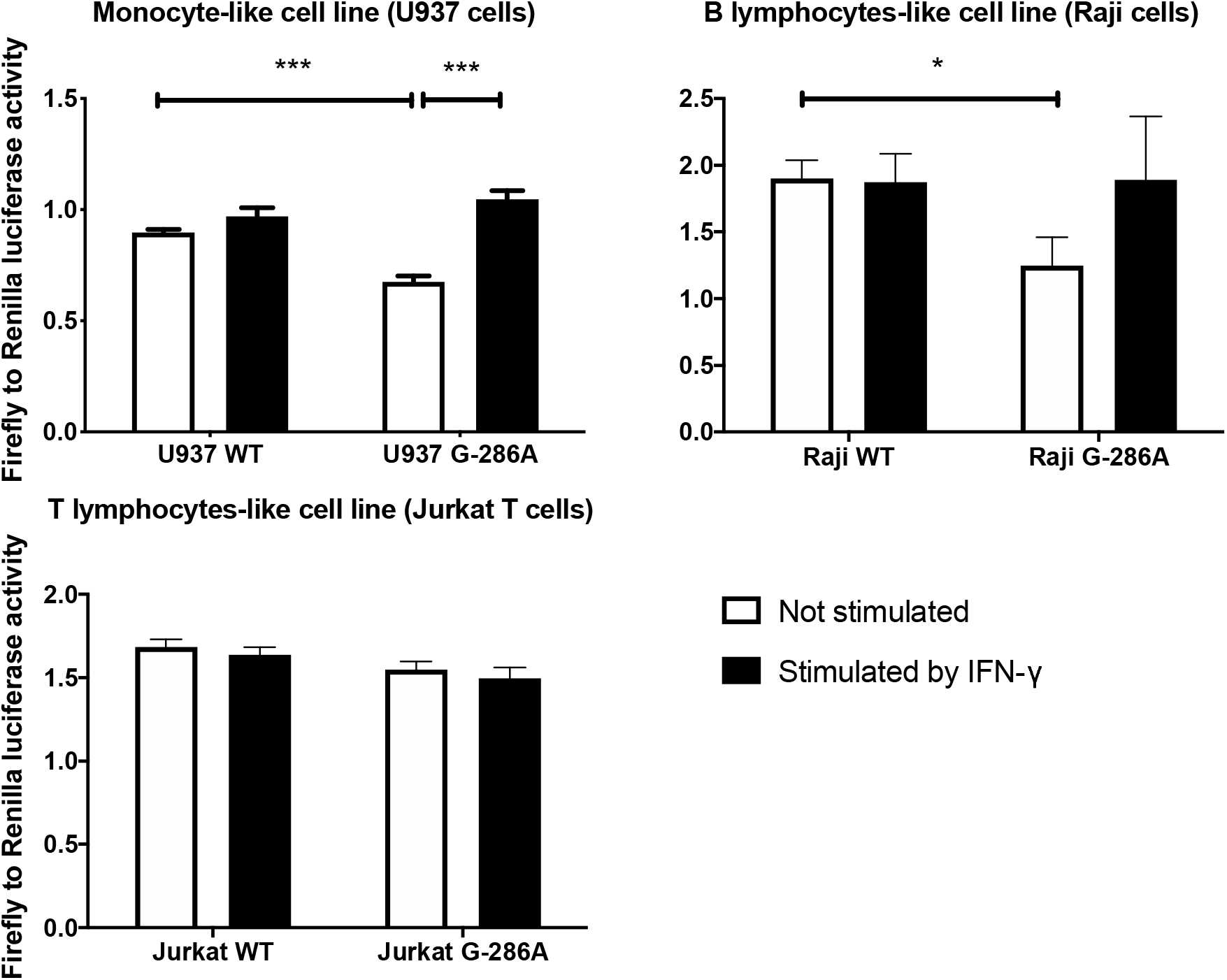
Transient transfection of U937, Raji and Jurkat T cells with CIITA-promoter-luciferase plasmid. Transfected cells were stimulated during 6h with IFN-γ. CIITA promoter III activity is represented by luciferase activity over empty vector. WT = Wild Type. P values were calculated using Mann-Whitney Test. ***P < 0.001 and*P < 0.05.

### Rs3087456 genotype effect on *CIITA* isoforms expression

To determine whether the *CIITA* rs3087456 polymorphism was associated with different expression of *CIITA* isoforms derived from the promoter pIII and pIV, mRNA of purified monocytes (CD14^+^) and B cells (CD19^+^) was quantified by Q-PCR. mRNA encoding *CIITA* pI isoform was not recovered in isolated monocytes and B cells from healthy donors (data not shown). In monocytes, no difference in basal expression level of pIII (*P* = 0.18) and pIV (*P* = 0.24) transcripts between AA or AG and GG genotypes (Figure 4). After 6 hours of *ex-vivo* cells stimulation with IFN-γ, pIII and pIV transcripts were significantly up-regulated (*P* < 0.0001, Figure 4) in monocytes for AA or AG as well as GG genotype (Figure 4). In B cells, no difference was observed in pIII or pIV transcripts expression in unstimulated cells. Following IFN-γ stimulation, only AA or AG genotype pIV transcripts expression significantly increased (*P* < 0.05, Figure 4). However, pIV transcripts induction following IFN-γ stimulation was much higher in monocytes (45 fold increase: from 1.46 [CI 95%, 0.59 to 2.28] to 66.98 [CI 95%, 38.32 to 164.6]) than in B cells (2 fold increase, from 4.40 [CI 95%, 2.37 to 8.27] to 8.70 (CI 95%, 4.42-12.05]) (Figure 4). Altogether rs3087456 polymorphism was not impacting *CIITA* isoforms basal expression, but is similarly inducible by IFN-γ in both AA or AG and GG in monocytes but not in B cells.

**Figure 4.**
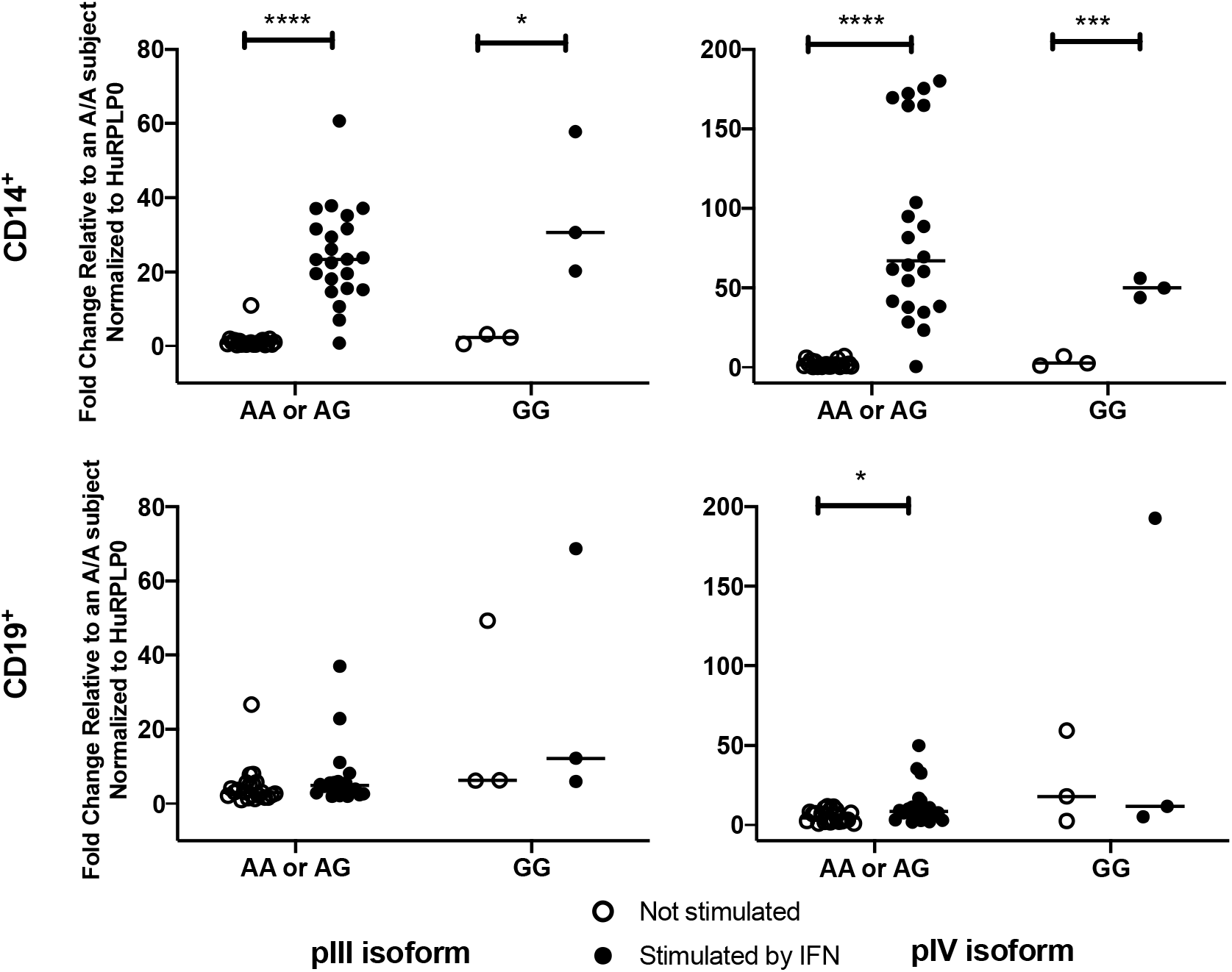
mRNA levels for promoter III (pIII) and promoter IV (pIV) isoforms. mRNA level in CD14^+^ (monocytes) and CD19^+^ (lymphocytes) from healthy volunteers. Cells were stimulated or not by IFN-γ during 6h before qPCR. Results are presented were calculated using Mann-Whitney Test. ****P < as median fold change relative to an AA subject normalized to HuRPLP0, and by genotype of CIITA rs3087456 in an A-dominant model. P values 0.0001, ***P < 0.001 and*P < 0.05.

## DISCUSSION

In this study, we showed that G-286A *CIITA* promoter polymorphism is associated with an increase 28-day mortality in septic shock and altered mHLA-DR kinetics. Monocyte HLA-DR impaired expression is related to an altered response to IFN-γ stimulation in G-286A. This regulatory effect seems to be related to altered basal pIII activity that is restored in G-286A after IFN-γ stimulation.

Gene polymorphisms is widely recognized as important determinants of host response to pathogens and infections have been a strong selective sieve of human genome evolution ^36,37^. In this respect, multiple human gene polymorphisms, positively selected in the population, are associated with survival during infection ^38,39^. For example, premature stop codon inactivating the caspase-12 gene has been suggested to provide a selective advantage through sepsis resistance in population that experience more infectious diseases ^40^. Nevertheless, different studies supported the idea that allele selected to resist to human pathogens, may play a role in autoimmune disease susceptibility ^41^. Recently, Vasseur et al ^31^ identified one selected SNP of *CIITA* promoter region, rs3087456, which is positively selected in humans and known to be associated with autoimmunity, and therefore may prove to be a strong candidate SNP in sepsis resistance. In our study, rs3087456 is associated with improved survival to septic shock confirming its potential selective function. In addition to Swanberg et al ^23^ showing that rs3087456 is correlated with MHC class II transcripts, we showed its correlation with expression of HLA-DR on the cell membrane of monocytes in patients with septic shock. In homozygous GG patients, our data showed that mHLA-DR remained low throughout the ICU stay, suggesting that persistent sepsis-induced immunodepression may be partially related to this polymorphism.

The effect of the rs3087456 polymorphism on the regulation of HLA-DR expression has not been previously studied. To our knowledge, this is the first study to investigate the regulatory effects of this promoter polymorphism. Our results suggest that GG genotype alters pIII activity in monocytes and B cells but not in T cells, in a cell-type specific manner. Unexpectedly, on basal condition (e.g. monocytes and B lymphocytes from healthy volunteers), pIII related transcripts induction is not significantly different between all haplotypes. Following *ex vivo* stimulation with IFN-γ, pIII transcripts expression was increased only in monocytes. Similarly, pIV transcripts expression is not different in the basal condition, but expression increases after IFN-γ only in monocytes. These suggest that, in basal condition (e.g. healthy), transcriptional induction counteracts altered promoter activity of GG or repressor inhibit transcription in AA or AG haplotypes, both hypotheses encompassing complex transcription regulatory mechanisms.

Few *CIITA* transcriptional regulatory mechanisms are described. Recently, immune-enhancing CpG and non-CpG oligodeoxynucleotides (ODNs) have been described as having a role in regulating expression of MHC class II and costimulatory molecules. Wang et al ^42^ showed that non-CpG ODNs (rODNs) down-regulate HLA-DR expression in monocytes and block promoter III-directed transcription of CIITA in these cells. The precise inhibitory molecular effects of rODN on pIII activity are not understood, and more specifically, its precise promoter interaction not studied. Lohsen et al ^43^ described a distal regulatory elements, known as hypersensitive site 1 - HSS1 (located ∼3kb upstream of pI) with PU.1, NF-κB, and Sp1 interacting with pIII through a looping interaction. Interestingly, proximal regulatory elements were described, and include inducers (e.g. AP-1, Sp1, CREB, NF-Y, ARE-1, ARE-2), and repressors (eg. PRDI-BF1/BLIMP-1, ZBTB32). Other mechanisms are involved in *CIITA* expression regulation. Among these, in B cells, restricted access of pI due to extensive DNA methylation induces a switch to pIII activation ^43^. The −300 region of *CIITA* has been well identified as an important proximal regulatory regions ^44^. It is plausible that pIII rs3087456 polymorphism may interfere with transcription in monocyte during sepsis and *CIITA* expression be rescued through IFN-γ stimulation. Whether this may be directly related to a cis- or trans-regulatory mechanism is unknown. Ni et al ^45^ identified a dependent distal enhancer regulating IFN-γ-induce expression of CIITA through pIV. This IFN-γ induce promoter activation is dependent on BRG1, an ATP-dependent remodeling factor, which promotes the formation of a dynamic three-dimensional chromatin loop and demonstrates the importance of distant enhancer/silencer regions in CIITA expression regulation. In our study we showed that expression in monocyte of pIV isoforms following IFN-γ is ∼2.7 fold higher than pIII, which confirms the preferential activation of pIV after IFN-γ induction. Altogether, these facts support the evidence that IFN-γ is a preferential inducer of CIITA expression that may overcome monocytic MHC class II phenotype triggered by rs3087456 polymorphism in septic patients. The detailed transcriptional mechanism is unknown but the complexity of the regulation of CIITA expression renders difficult the precise determination out of high-throughput promoter analysis.

Another observation of this study is the relation between *CIITA* promoter III SNP and mortality in septic shock. Outside the indirect relation between CIITA expression and mortality through mHLA-DR expression ^17,46,47^, there is increasing descriptions of the direct role of CIITA in regulating various effectors encountered during sepsis. CIITA has been shown to regulate the expression of several immune related genes such as interleukin (IL)-4, collagen α2, Fas ligand, and plexin A1. As an illustration, Nozell et al ^48^ showed that CIITA was able to inhibit MMP-9 expression, a matrix metalloproteinase involved in the degradation of extracellular matrix in lung injury. Interestingly, the inhibition of MMP-9 in a sepsis rat model with induced lung injury resulted in an increase survival ^49^. Hereby, we show that IFN-γ was able, in rs3087456 GG genotype, to restore *CIITA* promoter activity, CIITA pIII and pIV expression as well as HLA-DR expression in monocyte, suggesting that IFN-γ may be a suitable immunomodulatory therapy to recover MHC class II expression. In the seminal article by Döcke et al ^50^, authors showed that IFN-γ was able to restore monocyte functions in 8 patients with sepsis. More recently, in a cohort of 20 paediatrics and adults’ patients with sepsis, we showed that immunotherapy with IFN-γ was safe to improve the immune host defense, particularly the monocyte HLD-DR expression in patients with prolonged downregulation of mHLA-DR, and was associated with a very high survival rate ^51^. However, Leentjens et al ^52^ suggested in healthy volunteers with LPS-induced immunoparalysis that the modulatory effect of IFN-γ is variable, despite the significant increase in mHLA-DR expression. Interestingly, GM-CSF did not induce a significant increase in mHLA-DR in this study ^52^. This is in agreement with the study of Hornell et al ^53^ showing that GM-CSF increase CIITA type I and III transcript expression but not of type IV. The use of mHLA-DR to assess the risk of secondary infection after septic shock is well admitted ^14,15^. The use of a threshold of 8,000 AB/C to define a reduction in mHLA-DR expression to diagnose an acquired immunosupression gets more credence, in particular if it is lasting after day 3 post admission ^51,54^. Hereby, we showed that rs3087456 GG genotype was significantly associated with unresolving low mHLA-DR kinetics, predicting a persistent immunodepression phenotype in patients with septic shock.

This study opens two important perspectives: First, a genetic susceptibility affecting MHC class II master regulator CIITA is shown to be associated with defective mHLA-DR expression in patients with septic shock and dismal outcome. This allows to anticipate the development of sepsis-induced immunodepression. Second, the demonstration of IFN-γ Effect to rescue the functional consequence of this genotype, suggest the use of this genetic biomarker to identify patients who may benefit of IFN-γ-based immunomodulatory therapy. In conclusion, we report that *CIITA* rs3087456 GG genotype is associated with impaired m-HLA-DR expression in septic shock, predictive of development of persistent immunodepression, and demonstrate the effect of IFN-γ to rescue altered MHC class II expression.

## Data Availability

I declare that all the data mentioned in the manuscript are available.

## Acknowledgments

We thank Sergio Crovella (Department of Advanced Diagnostics, Institute for Maternal and Child Health-IRCCS ‘Burlo Garofolo’, Trieste, Italy and Department of Medical, Surgical and Health Sciences, University of Trieste, Trieste, Italy) for help with paper revisions.

## Contribution

ACL, DP enrolled the patients and collected clinical data

JM, MB, SH, KM, CB, VF performed experiments;

JM, MB, PT analyzed results and made the figures;

JM, MB, PT, DP, ACL designed the research;

JM, PT, MB wrote the paper;

All authors critically reviewed the manuscript.

