## Supplemental data for "CIITA G-286A promoter polymorphism impairs monocytes HLA-DR expression in septic shock and is rescued by interferon-γ"

**Supplementary File**

**Table 1. Primer sequences**

| **Assay** | **Name** | **Sequence** |
| --- | --- | --- |
| SNP genotyping | CIITA-P3 SNP-712 Taqman probe | ATGGGAGTCAGTATTATTTAGCATC[A/G]CTTTGGCGGGTCACCCCAAACCATC |
|  | CIITA-P3 SNP-286 Taqman probe | GAAGTGAAATTAATTTCAGAGGTGT[A/G]GGGAGGGCTTAAGGGAGTGTGGTAA |
| QPCR CIITA isoforms | Promoter1 f | CATGGTGGCAGCTCAC |
|  | Promoter3 f | CCCAAGGCAGCTCACA |
|  | Promoter4 f | GAACAGCGGCAGCTCA |
|  | Exon2 r | GTAGCCACCTTCTAGGG |
|  | HuRPLP03 f | AGGCTTTAGGTATCACCACTAA |
|  | HuRPLP03 r | ACATCACTCAGGATTTCAATGG |

Primer sequences are expressed as 5’ to 3’; f, forward primer; r, reverse primer.
